# Assessment of human milk samples obtained pre- and post-influenza vaccination reveals a poor boosting of seasonally-relevant, hemagglutinin-specific antibodies

**DOI:** 10.1101/2023.01.30.23285124

**Authors:** Xiaoqi Yang, Claire DeCarlo, Alisa Fox, Nicole Pineda, Rebecca L.R. Powell

## Abstract

Influenza (flu) vaccination prevented over 100,000 hospitalizations and 7000 deaths from flu over the 2019-2020 season in the USA [1]. Infants <6 months are the most likely to die from flu, though flu vaccines are only licensed for infants >6□months old. Therefore, it is recommended that flu vaccination occur during pregnancy, as this reduces severe complications; however, vaccination rates are suboptimal, and vaccination is also recommended postpartum [2-6]. For breast/chest-fed infants, the vaccine is believed to elicit protective and robust seasonally-specific milk antibody (Ab) [4, 7]. Few comprehensive studies exist examining Ab responses in milk after vaccination, with none measuring secretory Ab (sAb). Determining whether sAbs are elicited is critical, as this Ab class is highly stable in milk and mucosae [8, 9]. In the present study, our aim was to determine to what extent specific Ab titers in the milk of lactating people were boosted after seasonal influenza vaccination. Over the 2019-2020 and 2020-2021 seasons, milk was obtained pre- and post-vaccination and assessed for specific IgA, IgG, and sAb against relevant hemagglutinin (HA) antigens by a Luminex immunoassay. IgA and sAb were not found to be significantly boosted, while only IgG titers against B/Phuket/3073/2013, included in vaccines since 2015, exhibited an increase. Across the 7 immunogens examined, as many as 54% of samples exhibited no sAb boost. No significant differences for IgA, sAb, or IgG boosting were measured between seasonally-matched versus mismatched milk groups, indicating boosting was not seasonally-specific. No correlations between IgA and sAb increases were found for 6/8 HA antigens. No boost in IgG-or IgA-mediated neutralization post vaccination was observed. This study highlights the critical need to redesign influenza vaccines with the lactating population in mind, wherein the aim should be to elicit a potent seasonally-specific sAb response in milk. As such, this population must be included in clinical studies.

## Introduction

Influenza virus circulates globally causing annual outbreaks in the winter months of a given geographic area (seasonal flu), though this is less the case in warmer climates which might experience multiple peaks or consistent levels of infection [10]. The flu virus attachment protein hemagglutinin (HA) utilizes sialic acid on the epithelial cell surface, primarily in the nose, throat, and lungs of mammalian hosts [11, 12]. Orthomyxoviridae viruses possess an RNA genome and utilize an RNA-dependent RNA polymerase for genomic replication. As these polymerases lack a proofreading capacity, mutation during replication is expected, causing antigenic drift. Over time, 4 influenza species have emerged, influenza A-D. Influenza A and B are far more virulent and common, and all licensed flu vaccines protect against these species. Within flu A, 18 HA and 11 NA subtypes have been identified, though only a minority of these HA and NA subtypes are present in viruses that circulate among humans [13]. Due to the segmented flu genome, reassortment of these segments can occur during superinfection, causing a rapid antigenic shift. Both of these mechanisms, particularly the latter, results in the need for seasonal flu vaccines which are only considered effective for one or possibly a few years depending on the circulating flu strains [12].

Flu typically causes fever and respiratory symptoms that resolve without serious intervention within 1-2 weeks; however ∼3-5 million cases of severe illness and 250,000– 500,000 deaths from flu occur globally each year [10]. Leading causes of death from flu include primary and secondary (bacterial) pneumonia, and exacerbation of pre-existing pulmonary conditions. Licensed flu vaccines are highly effective at preventing the complications of flu infection [1]. Several egg- and cell-based inactivated flu whole virus and recombinant vaccines are approved by the FDA to be given by intramuscular (IM) injection; these are largely quadrivalent formulations [14]. During the 2019–2020 flu season, it is estimated that vaccination prevented 7.09 million illnesses, 3.46 million medical visits, 100,000 hospitalizations and 7,100 deaths associated with flu [1]. Vaccination against flu has been shown to reduce the risk of a child’s need for admission to the intensive care unit by 74%, the risk of flu-associated death by 51% and 65% for high-risk and otherwise healthy children, respectively [15].

Flu can cause significant respiratory illness among infants, accounting for ∼3000 hospitalizations per year in the United States, with infants <6 months being the most likely to die from flu compared to any age group [16, 17]. Flu vaccines are only licensed for infants >6□months old, in part due to maternal IgG transferred *in utero* that is likely to suppress the infant’s response to the vaccine before this time [18, 19]. Therefore, it is recommended that flu vaccination occur during pregnancy when it is seasonally relevant, particularly during the 3^rd^ trimester, as placental transfer of antibody (Ab) is maximal at this time [20, 21]. Various studies have demonstrated that vaccination during pregnancy is effective at reducing infant infection, respiratory illness, and severe respiratory complications [4-6]. Despite this recommendation and the knowledge that the flu vaccine is safe and effective for pregnant and lactating women, vaccination rates are suboptimal [2, 3]. Furthermore, depending on the timing of the pregnancy relative to the flu season, vaccination during pregnancy may not be possible. Consequently, vaccinating against flu is recommended postpartum to provide a cocooning effect for the baby, but also because for breast/chest-fed infants, the vaccine is believed to boost existing milk Ab and elicit a robust seasonally-specific milk Ab response that is likely protective [4, 7].

Human milk contains approximately 0.7g/L IgA, which comprises ∼90% of the total immunoglobulin (Ig) in milk [9, 22]. Approximately 2% of Ig in milk is IgG, as humans rely on placentally transferred IgG for systemic immunity [9]. Milk-derived Abs are likely most active in the oral cavity and upper GI tract of infants, though ∼30%-50% of these Abs have been shown to resist degradation in the stomach for as long as 2h suggesting that they are functional throughout the GI tract and possibly beyond [23]. Milk IgG is derived primarily from the serum with a minority arising from local mammary production. B cells in the mammary gland that ultimately produce IgA (and to a lesser extent, IgM) that becomes secretory (s)IgA predominately originate from the gut-associated lymphoid tissue (GALT), exemplifying a critical *entero-mammry* link, wherein the sAbs found in human milk echo the immunogens identified in the maternal GI tract (and airways) [24, 25]. This IgA is polymerized (mostly dimerized) with a joining (J) chain within the B cell prior to secretion, and then bound by the polymeric Ig receptor (pIgR) on mammary epithelial cells. PIgR is cleaved as it transports Ab into the milk, leaving the secretory component (SC) attached and resulting in sAb [24, 26].

Relatively few comprehensive studies exist examining the Ab response in milk after vaccination. The few studies that have examined the milk Ab response after influenza, pertussis, meningococcal and pneumococcal vaccination have generally found specific IgG and/or IgA that tends to mirror the serum Ab response, though none of these studies measured secretory Ab or determined if sIgA was elicited, and data regarding the protective capacity of these milk Abs is conflicting or confounded by the effects of placentally-transferred Ab [4, 27-33]. Determining whether or not sAbs are elicited in milk after infection or vaccination is critical, as this Ab class is highly stable and resistant to enzymatic degradation in all mucosae - not only in the infant oral/nasal cavity, but in the airways and GI tract as well [8, 9]. Vaccine-elicited Abs in human milk may be protective for the infant; remarkably, studies addressing this response in women immunized postpartum are lacking [4, 7]. The present study aims to fill the knowledge gap concerning whether vaccination of lactating women elicits Abs in their milk that is likely to be protective against seasonal influenza strains.

## Material and methods

### Study participants and milk samples

This study was approved by the Institutional Review Board (IRB) at Mount Sinai Hospital (IRB 19-01243). Individuals were eligible to have their milk samples included in this analysis if they were lactating and were planning to receive a seasonal influenza vaccine.

Participants were excluded if they had any acute or chronic health conditions affecting the immune system. Participants were recruited locally in NYC via social media during the 2019-2020 and 2020-2021 flu seasons, and subject to an informed consent process. All demographic information on participant milk samples is shown in Table 1. Sample IDs are known only within the research group. Given the diversity of participant ages and stages of lactation, this study sample can be considered representative of a larger population.

**Table 1:** Study Participant Information.

| **Participant ID\*** | **Race** | **Ethnicity** | **Participant Age (years)** | **Months Postpartum** |
| --- | --- | --- | --- | --- |
| FLU100 | White | Not Hispanic or Latino | 30-35 | 7 |
| FLU102 | White | Hispanic or Latino | 30-35 | 6 |
| FLU103 | White | Hispanic or Latino | 20-25 | 5 |
| FLU104 | White | Not Hispanic or Latino | 41-45 | 23 |
| FLU113 | White | Hispanic or Latino | N/A | 4 |
| FLU114 | White | Hispanic or Latino | N/A | N/A |
| FLU115 | White | Hispanic or Latino | N/A | N/A |
| FLU116 | Asian | Not Hispanic or Latino | 41-45 | 3 |
| FLU118 | White | Not Hispanic or Latino | 30-35 | 6 |
| FLU119 | Two or more races | Not Hispanic or Latino | N/A | N/A |
| FLU120 | Asian | Not Hispanic or Latino | N/A | N/A |
| FLU121 | Asian | Not Hispanic or Latino | N/A | 11 |
| FLU122 | Two or more races | Not Hispanic or Latino | N/A | N/A |
| FLU200 | White | Hispanic or Latino | 26-29 | 11 |
| FLU201 | White | Not Hispanic or Latino | 30-35 | 6 |
| FLU202 | White | Not Hispanic or Latino | 30-34 | 6 |
| FLU203 | White | Not Hispanic or Latino | 30-34 | 43 |
| FLU204 | White | Not Hispanic or Latino | 30-35 | 5 |
| FLU205 | White | Not Hispanic or Latino | 36-40 | 6 |
| FLU206 | White | Not Hispanic or Latino | N/A | 11 |
| FLU207 | White | Not Hispanic or Latino | 30-35 | 8 |
| FLU208 | White | Not Hispanic or Latino | 36-40 | 5 |
| FLU209 | White | Not Hispanic or Latino | 41-45 | 19 |
| FLU210 | White | Not Hispanic or Latino | 30-35 | 9 |
| FLU211 | White | Not Hispanic or Latino | 41-45 | 22 |
| FLU212 | Asian | Not Hispanic or Latino | N/A | 11 |

\*Sample IDs are known only within the research group

Participants were asked to collect approximately 75-100mL of milk per sample into a clean container using electronic or manual pumps, just prior to, and 2 weeks after receiving a seasonal influenza vaccine. All participants received a quadrivalent intramuscular (IM) vaccine in NYC. Milk was frozen in participants’ home freezers until samples were picked up, and stored at -80°C until processing and testing. Milk samples were then thawed, centrifuged at 800g for 15 min at room temperature, fat was removed, and the de-fatted milk transferred to a new tube. Centrifugation was repeated 2x to ensure removal of all cells and fat.

### Luminex Binding Assay

Influenza HA antigens were produced as previously described and covalently coupled individually to magnetic beads using a two-step carbodiimide reaction with the xMAP Ab Coupling (AbC) Kit according to manufacturers’ instructions (Luminex, Austin, TX) [34]. Carboxylated xMAP beads were coupled at 1μg protein/million beads (A/Brisbane/02/2018 (H1N1) pdm09-like virus, A/Kansas/14/2017 (H3N2)-like virus, B/Colorado/06/2017-like virus (B/Victoria lineage), B/Phuket/3073/2013-like virus (B/Yamagata lineage), A/Guangdong-Maonan/SWL1536/2019 (H1N1) pdm09-like virus, A/Hong Kong/2671/2019 (H3N2)-like virus) or 5μg protein/million beads (B/Washington/02/2019-like virus (B/Victoria lineage)) in an optimized buffer comprised of 0.5% Casein (Thermo Scientific) and 0.05% sodium azide (Ricca chemical) in PBS. The coupled beads were counted, diluted in buffer to a concentration of 500,000 beads/mL and stored at 4°C for up to 1 month prior to use. Each antigen was bound to a bead with a different fluorescent profile. Fluorescence was measured using a Luminex FlexMAP3D device with xPONENT 4.2 software. Samples were serially diluted and tested in duplicate according to manufacturer’s protocol, using 2ug/mL biotinylated goat anti-human-IgA or –IgG (Fisher), or 2ug/mL biotinylated sheep anti-human-secretory component (MuBio) diluted in buffer.

Results are shown as mean fluorescent intensity (MFI). Wells with no milk served as background controls and values from these wells were used to ensure reproducibility and subtracted from test well MFIs. Positive control milk pools were also used to ensure reproducibility. After background subtraction, MFI values at each dilution were logarithmically transformed and fit to 4-parameter curves. Curves were used to interpolate endpoint titers at a cutoff MFI of 200. Mann-Whitney U tests were used to assess significant differences between unpaired grouped data pre- and post-vaccination. Correlation analyses were performed using Spearman correlations. Data was analyzed in GraphPad Prism, with all statistical tests being 2-tailed and significance level set at p-values < 0.05.

### IgA and IgG extraction

Total IgA or IgG was extracted from 25 – 100mL of milk or 1mL of plasma using peptide M or protein A/G agarose beads (Thermo), respectively, following manufacturer’s protocols. Ig was concentrated using Amicon Ultra centrifugal filters (10 kDa cutoff; Fisher) following manufacturer’s protocol, and quantified by Nanodrop.

### Influenza microneutralization assay

Assay was adapted from that described in [35]. Flat-bottom 96-well cell tissue culture plates (Corning Costar) were seeded in 100µl of UltraMDCK (EMEM) culture medium containing 1.8 × 10^5^ MDCK cells and incubated at 37°C overnight. The following day, RDE-treated serum samples diluted to 1:7 were 3-fold serially diluted in UltraMDCK (EMEM) containing TPCK-treated trypsin (1ug/ml, infection medium) in a separate 96-well tissue culture plate. Titrated milk or plasma Ab were incubated with influenza A/Brisbane/02/2018 (H1N1) pdm09-like virus (22.2ng/ml) for 1h at room temperature with shaking. Cell plates were washed with PBS, and 100 µl of the virus-Ab mixture was added. After a 1h incubation at 33°C the mixture was removed, cells were washed with PBS, and 50 µl of titrated Ab plus 50 µl infection medium were added to each well. The cells were then incubated at 33°C for 48 h. To measure microneutralization activity, 50 µl of the supernatant was transferred into a separate V-bottom 96-well plate, and 50 µl of 0.5% chicken red blood cells in PBS were added. The samples were read after 45 min of incubation at room temperature.

## Results

Thirteen study participant milk sample pairs per influenza season were assessed for specific IgA, IgG, and sAb binding against each relevant HA strain included in either the 2019-2020 or 2020-2021 seasonal influenza vaccines using a 4-plex Luminex-based immunoassay. Milk was processed as described in methods, and skimmed acellular milk was serially diluted to generate MFI curves in duplicate for each experiment (SFig. 1). It was found that across both seasons, all milk samples pre- and post-vaccination exhibited binding to all HA antigens tested (i.e., MFI at the lowest dilution tested of 1/2 > MFI of the ‘no milk’ control used for background subtraction); however, IgA, IgG, and sAb reactivity to B/Washington/02/2019 was significantly lower than to those of the other 2020-2021 antigens (p<0.0094; SFig. 1), even when coated to Luminex beads at 5x concentration.

Titration curves were used to determine endpoint binding milk titers at an MFI cutoff value of 200. Geometric mean IgA, sAb, and IgG endpoint titers for pre- and post-vaccination sample groups against each antigen for each season were determined and compared. As noted above, mean IgA, sAb, and IgG endpoint titers against B/Washington were significantly lower compared to all other antigens (p<0.0001; Fig. 1a). For the remaining antigens, mean IgA endpoint titers ranged from 111 – 528 and were not found to be significantly boosted comparing pre-to post-vaccination levels for either season studied (Fig. 1a). Mean sAb endpoint titers (excluding B/Washington) ranged from 7.1 – 154 and were also not found to be significantly boosted comparing pre-to post-vaccination levels for either season studied (Fig. 1a). Mean IgG endpoint titers (excluding B/Washington) ranged from 16 – 134, with post-vaccination titers against B/Phuket/3073/2013 in the 2019 – 2020 season exhibiting a significant increase compared to pre-vaccination levels (p=0.0387; Fig. 1a). IgG reactivity against all other antigens was not significantly increased in either season.

**Figure 1:**
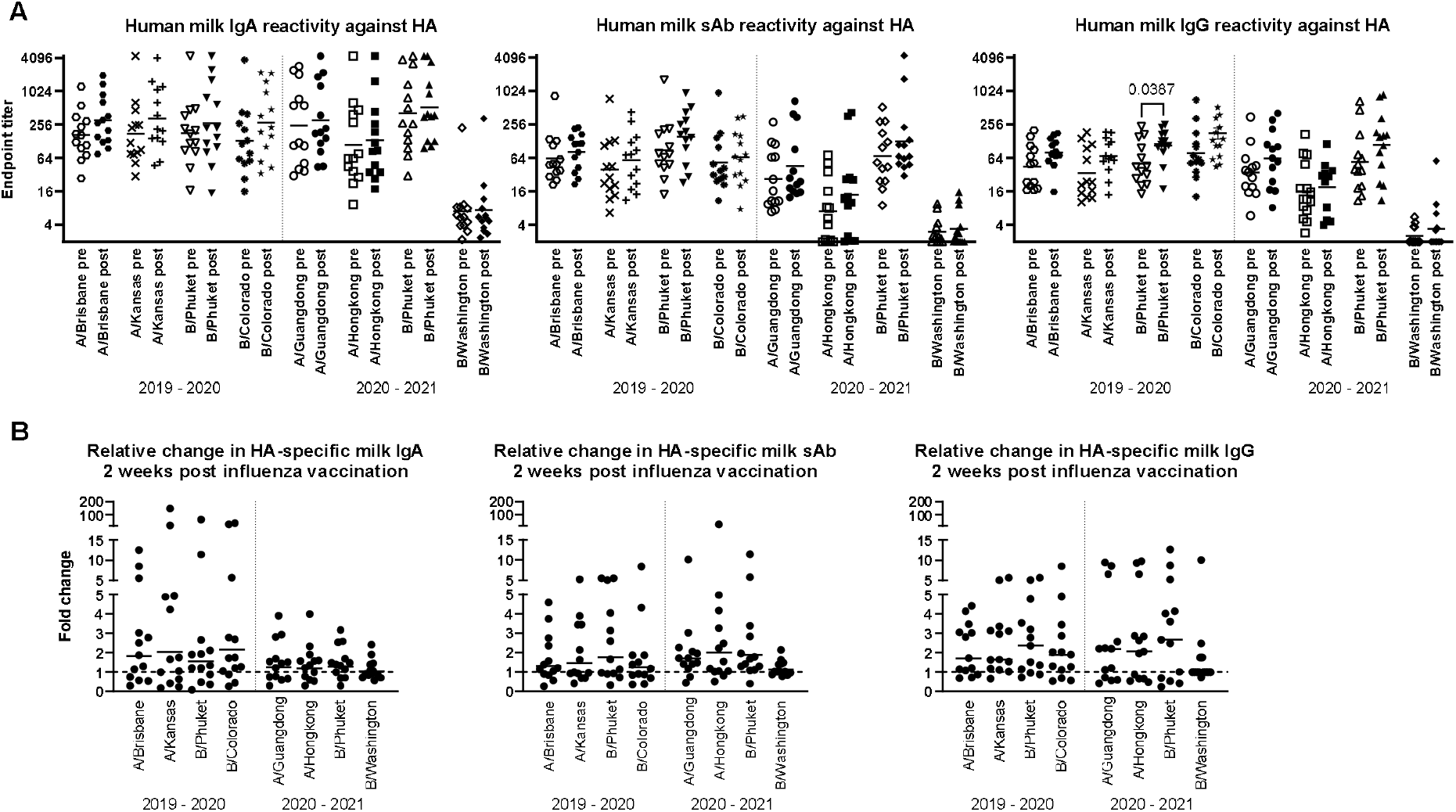
Human milk antibody titers to HA post influenza vaccination demonstrate a poor boosting effect. Participant milk sample pairs were assessed for specific IgA, IgG, and sAb binding against each relevant HA strain using a 4-plex Luminex-based immunoassay. Skimmed acellular milk was serially diluted to generate MFI curves in duplicate for each experiment. After background subtraction, MFI values at each dilution were logarithmically transformed and fit to 4-parameter curves. Curves were used to interpolate endpoint titers at a cutoff MFI of 200. Mann-Whitney U tests were used to assess significant differences between unpaired grouped data pre- and post-vaccination. (a) IgA, sAb, and IgG titers measured pre- and post-vaccination against each HA included in each seasonal vaccine are shown. (b) Fold changes in IgA, sAb, and IgG titers are shown. Dotted line at 1 indicates no change.

The fold change of pre-to post-vaccination endpoint titers against each antigen was determined (Fig. 1b). It was found that for IgA, the geometric mean fold titer change after vaccination in the 2019-2020 season was 1.56 – 2.14 depending on the antigen tested, with 31% - 46% of samples exhibiting no change or a decrease in titer after vaccination. Notably, 15% - 38% of samples exhibited a >4-fold increase in IgA titer (Fig. 1b). In the 2020-2021 season, vaccinees exhibited a geometric mean 1.04 – 1.27-fold increase in specific milk IgA titers, with 31% - 38% of samples exhibiting no change or a decrease in specific titers (excluding B/Washington), and no ‘high’ titer group being evident as seen in the previous season. For sAb, the geometric mean fold titer change after vaccination in the 2019-2020 season was 1.24 – 1.77, ranging up to an 8.4-fold increase, and 38% - 54% of samples exhibiting no change or a decrease in sAb titer (Fig. 1b). In the 2020-2021 season, mean specific sAb titers increased 1.13 – 1.98-fold, ranging up to a 26-fold increase, and 7% - 38% of samples exhibiting no change or a decrease in sAb titer. Finally, it was found that specific IgG titers after vaccination in the 2019-2020 season exhibited a mean fold increase of 1.73 – 2.09, ranging up to an 8.5-fold increase, with 15% - 23% of samples exhibiting no change or a decrease in specific IgG. In the 2020-2021 season, mean specific IgG titers increased 1.36 – 2.01-fold, ranging up to a 12.7-fold increase, with 31% - 38% of samples exhibiting no change or a decrease in specific IgG titer (Fig. 1b).

The milk samples obtained from the 2019-2020 cohort were also assessed for Ab reactivity against the 2020-2021 influenza season HA antigens as a measure of general/cross-reactive anti-HA boosting after seasonal influenza vaccination as opposed to a seasonally-specific HA Ab response (i.e., a ‘mismatched’ Ab response; SFig. 4). As above, fold changes in endpoint binding titers from pre-to post-vaccination samples were determined. These fold changes were compared to the ‘matched’ fold changes detailed above, against the 3 relevant HA antigens (A/Guangdong-Maonan/SWL1536/2019, A/Hong Kong/2671/2019, and B/Washington/02/2019 [B/Phuket/3073/2013 was included in both seasons’ vaccines and therefore not included in this analysis]). It was evident that after vaccination, there were no significant differences between the fold change in Ab titers measured in seasonally-matched versus mismatched milk samples, against any 2020 – 2021 HA antigen for IgA, sAb, or IgG (Fig. 2).

**Figure 2:**
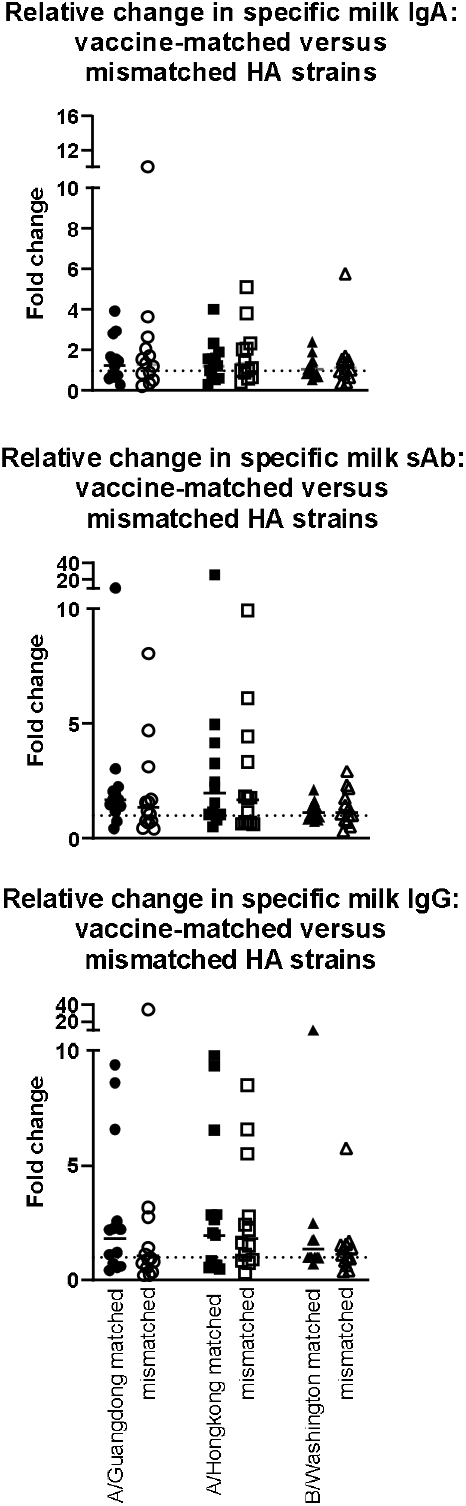
Milk antibody titers against mismatched HA immunogens indicate boosting effect is not seasonally-specific. Assays were performed as in Fig. 1. Milk samples obtained in the 2019-2020 season were tested against 2020-2021 HA antigens. As B/Phuket was included in both seasons’ vaccines, this HA was not tested. IgA, sAb, and IgG fold changes are shown separately.

Additionally, IgA and sAb fold increases from pre-to post-vaccine samples against each seasonally-matched antigen were used in Spearman correlation analyses as a measure of the sIgA versus monomeric (serum-derived) IgA response. For the 2019 – 2020 season, no correlation between IgA and sAb increases after vaccination were found for the response against A/Brisbane, A/Kansas, or B/Phuket, while a moderate positive correlation was observed for the response against B/Colorado (r=0.77; p=0.0125; Fig. 3). For the 2020 – 2021 season, no correlation between IgA and sAb increases after vaccination were found for the response against A/Hong Kong, B/Phuket, or B/Washington, while a moderate positive correlation was observed for the response against A/Guangdong (r=0.75; p=0.0098).

**Figure 3:**
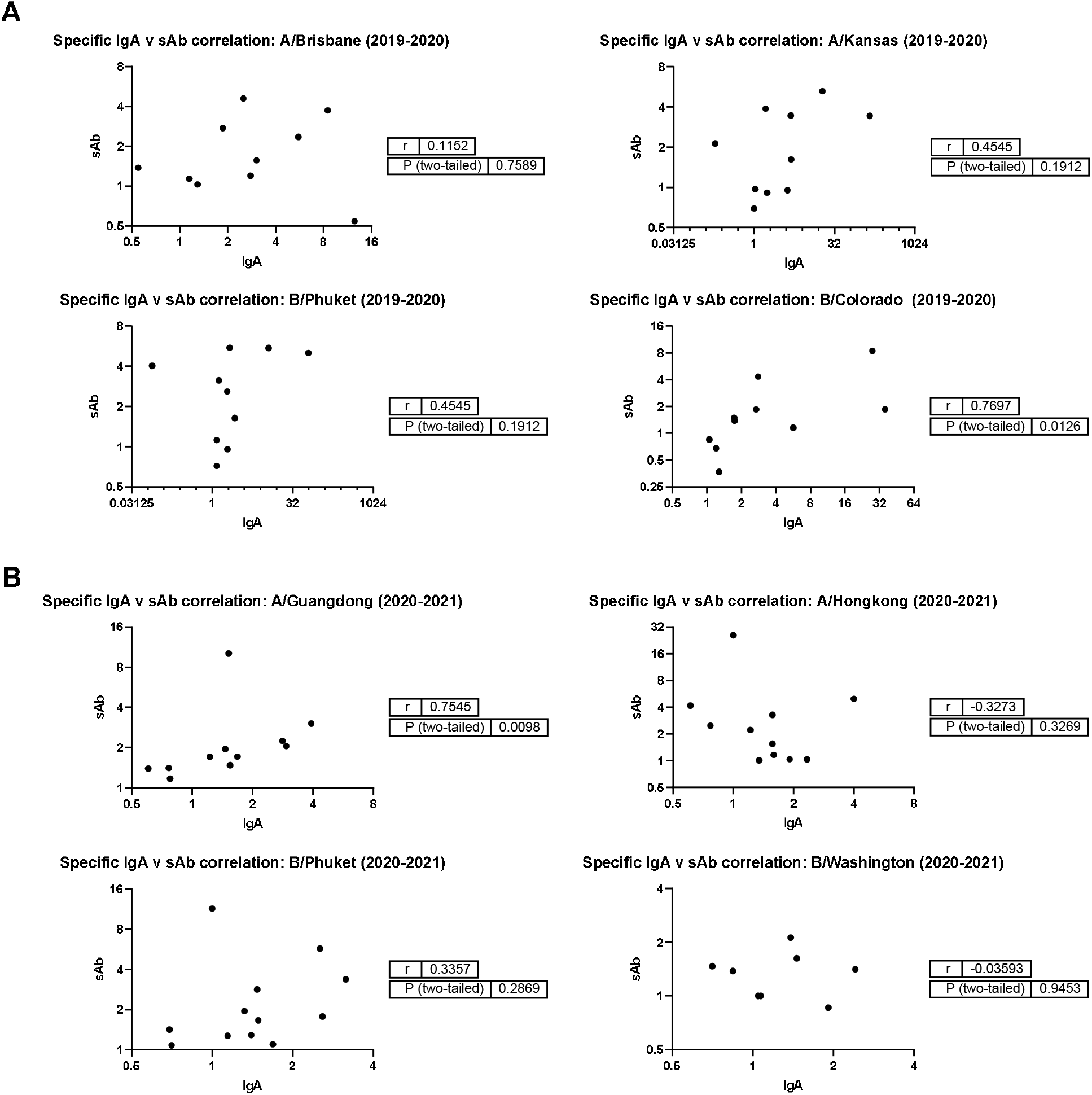
Magnitudes of IgA and sAb boosting post-vaccination do not tend to correlate, indicating most boosted IgA is not in secretory form. Any sample pairs found to exhibit an IgA and/or sAb titer increase in Fig. 1b were included in a Spearman correlation analysis to determine if the fold increases were related. (a) 2019-2020 season. (b) 2020-2021 season.

The capacity for milk-derived IgG and IgA to neutralize influenza virus infection was measured as described in [35]. This assay differs from the hemagglutination inhibition assay (HAI) in that the amount of input virus is not sufficient to show hemagglutination without virus replication, and therefore virus replication is required for observable agglutination of the red blood cells by the virus. While Abs that show activity in the HAI will almost always neutralize, some Abs do not show HAI but still can inhibit virus infection [35]. Therefore this assay more accurately measures the ability of Abs to neutralize viral replication. Purified, total IgG and IgA from milk samples exhibiting a fold increase in Ab titer by Luminex assay above were titrated and the neutralization capacity measured. Purified positive control plasma IgG inhibited hemagglutination, exhibiting a mean neutralization endpoint concentration of 25.4ug/mL. However, most milk Ab samples tested exhibited no neutralization even at the highest concentration used (500ug/mL). One IgA sample, FLU120, exhibited neutralization activity pre-vaccination (endpoint concentration, 1.23ug/mL), which increased post-vaccination (endpoint concentration, 0.13ug/mL; Fig. 4). FLU113 IgA did not neutralize pre-vaccination but did exhibit modest neutralization activity post-vaccination (endpoint concentration, 89ug/mL; Fig. 4). These 2 samples also demonstrated neutralizing IgG activity. FLU120 IgG was found to exhibit a neutralization endpoint concentration of 0.05ug/mL pre-vaccination, with this activity decreasing post-vaccination (endpoint concentration, 0.15ug/mL). FLU113 IgG neutralization activity was measurable pre-vaccination (endpoint concentration, 0.41ug/mL), and increased post-vaccination (endpoint concentration, 0.13ug/mL; Fig. 4). Overall, no significant differences were found in IgG-or IgA-mediated influenza neutralization activity between the pre-vaccination and post-vaccination groups (Fig. 4).

**Figure 4:**
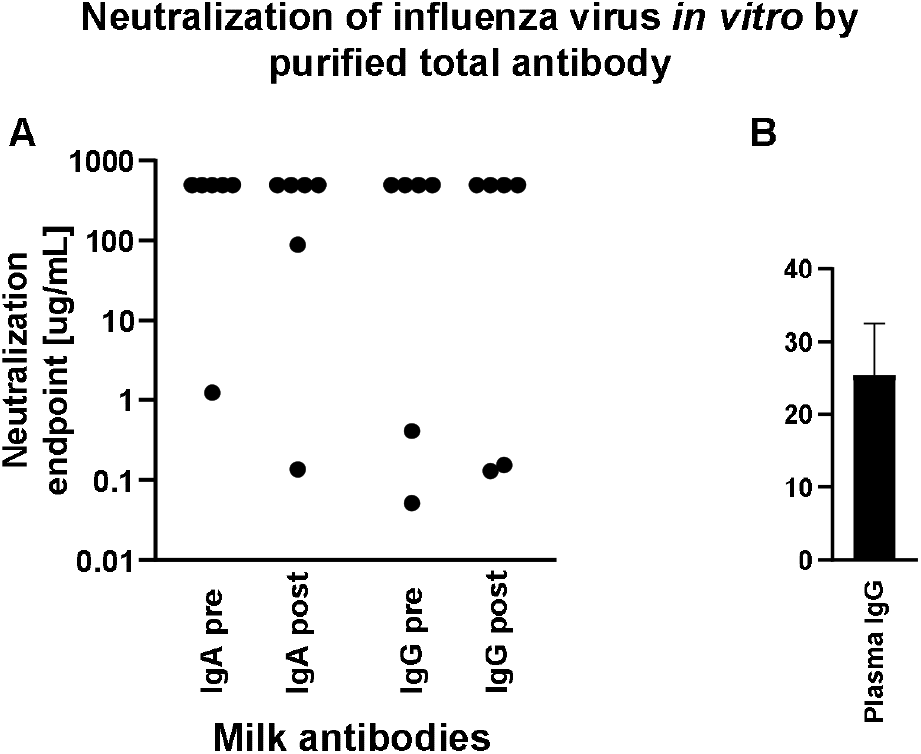
Purified milk IgA and IgG exhibit poor influenza neutralization and no difference was observed comparing pre-vs post-vaccination samples. Total IgA or IgG was extracted from milk or control plasma using peptide M or protein A/G agarose beads. Influenza microneutralization assay was adapted from that described in [35] using MDCK cells and A/Brisbane/02/2018 (H1N1) pdm09-like virus. Neutralization endpoint was defined as the lowest concentration of milk IgA/IgG or plasma IgG shown to exhibit neutralization. Samples were tested in duplicate. (a) Purified milk IgA and IgG from 6 participants pre- and post-vaccination. Note that those with an endpoint titer of 500ug/mL (the highest concentration tested) exhibited no neutralization. (b) Purified plasma IgG. This individual was not one of the milk donors but had a history of seasonal flu vaccination.

## Discussion

In the present study, our aim was to determine to what extent specific Ab titers in the milk of lactating people were boosted after seasonal influenza vaccination. Though a small number of studies examining the human milk An response to influenza vaccines have been done, which find measurable titers to the HA immunogens, and have strongly suggested milk Ab against HA is protective for breast/chest-fed infants over the course of lactation, these studies have not specifically examined true specificity of the response against seasonal HA antigens, nor have they examined baseline vs. post-vaccine boosting. Herein we have determined that although all samples exhibited measurable IgG, sAb, and IgG HA titers, these titers were not significantly boosted by vaccination, with the exception of IgG titers against one HA immunogen, B/Phuket, which notably, has been included in seasonal influenza vaccine in the Northern hemisphere since 2015 and therefore it is likely that individuals exhibit heightened reactivity to this HA due to its reoccurring inclusion in the seasonal vaccine and/or increased likelihood of infection with this strain over time. Given that our recent study of the milk Ab response to COVID-19 vaccines have shown IM vaccination to elicit an IgG-dominant response in milk, it is not surprising that the only significant indication of a boosting effect after influenza vaccination was observed for IgG [36]. Individually examining the relative fold changes from baseline to post-vaccine titers of HA-specific Ab in milk, it was clear that there was only a very minimal effect for most participants, wherein hardly a 2-fold titer increase was observed for any class of Ab against any HA examined. For many samples, no increase was observed at all. Intriguingly, in the 2019-2020 season, a clear subset of samples exhibited a notably higher fold-increase in specific IgA, while this was not observed for the 2020-2021 season. This difference may likely have been the result of more frequent infection among the participants for the 2019-2020 season, prior to their baseline samples. Though specific information on influenza infection for these participants is not known, it is well established that relative the 2020-2021 season, which was significantly affected by COVID-19 pandemic-induced masking and social distancing behaviors, the 2019-2020 season was more severe in terms of infection rate [37]. Notably, we determined that for all relevant HA antigens, seasonally-matched versus mismatched milk samples exhibited no difference in extent of specific boosting for IgA, sAb, or IgG, indicating that any measured titer increases were not seasonally-specific.

Presently we also measured correlation between IgA and sAb responses in milk after vaccination as a measure of the extent of the sIgA response versus monoclonal IgA from serum. We found that in general the responses did not correlate, with the exception of a single immunogen in each season, which is again likely a reflection of infection history more than any vaccine-induced effect. Our recent work analyzing the milk Ab response to COVID-19 vaccines has provided the unique opportunity to measure milk Ab in the absence of pre-existing immunity to the immunogen, which cannot be done with influenza; therefore, all vaccine data assessed in the current study is influenced by the particular influenza vaccination and infection history of each study participant [36]. Our COVID study clearly demonstrated that without the extremely potent serum Ab response induced by mRNA-based vaccination, it can be expected that only a relatively low or absent titer will be observed in milk [36]. Follow-up studies have also shown that these responses are short-lived [38]. This is in striking contrast to the infection response, which exhibits a classical mucosal profile that is sIgA-dominant and long-lasting. It is evident that IM vaccines, especially those using more ‘traditional’ vaccine platforms, are far less than ideal for eliciting a robust milk Ab response, which not only should be high-titer, but also, rich in sAb. It is reasonable to hypothesize that a mucosal vaccine would be preferable to induce this response. One comparative study found that in humans, live-attenuated influenza vaccine, which is administered as a nasal spray, actually elicited lower levels of serum influenza-specific Ab compared to an inactivated IM vaccine, but significantly greater levels of IgA in the nasal mucosa [39]. Based on these data, a separate study demonstrated that for IgA, milk titers were equivalent or lower for nasal spray recipients compared to IM vaccine, and for milk IgG, titers were significantly lower for nasal spray recipients, generally mirroring the serum response [7]. Notably this study did not measure sAb, nor did it report baseline milk titers. It is evident that the intranasal route is not likely to induce a potent milk sAb response.

Individuals who receive the influenza vaccine postpartum do not transmit seasonally-relevant IgG to the fetus in utero. These babies are therefore reliant on passive protection from milk Ab. One of the few studies measuring the milk Ab response to influenza vaccine examined women vaccinated in the 3^rd^ trimester [4]. This study demonstrated that influenza-specific IgA levels in milk were significantly higher in influenza vaccinees than controls for 6 months after birth, though neutralizing Ab in milk was only higher at birth and not at subsequent time points [4]. It is difficult to compare this study, which was performed in Bangladesh, to our present study, though it is highly likely that the Bangladeshi women had never received a previous influenza vaccine and that their infection history was significantly varied compared to our participants. Importantly, the Bangladeshi study did show that breastfeeding in the first 6 months of life significantly decreased respiratory illnesses in infants of mothers in the influenza group compared to those of pneumococcal-vaccinated controls [4]. The impact of vaccination occurring during pregnancy and the concomitant transfer of maternal IgG in utero in this study is unknown.

In conclusion, this study demonstrated that at least in the context of urban NYC participants, seasonal influenza vaccination elicits a poor boosting of specific milk Ab. These data should serve to strengthen the campaign to vaccinate pregnant people against influenza, at any time of year so as to achieve greater coverage, as well as the campaigns promoting vaccine safety for this population and efficacy of reducing infant morbidity and mortality. Even more critically, this study, as well as our published work examining the milk Ab response to COVID-19 vaccines, highlight the critical need to design influenza vaccines – and all other vaccines – with the lactating population and the breast/chest-fed infant in mind, and the importance of including this population in clinical studies.

## Data Availability

All data produced in the present study are available upon reasonable request to the authors

## Acknowledgements

We are indebted to our study participants. We thank Raffael Nachbagauer and Meagan McMahon for technical assistance and influenza reagents. This study was funded by the Icahn School of Medicine at Mount Sinai.

---

**Supplemental Figure 1:**
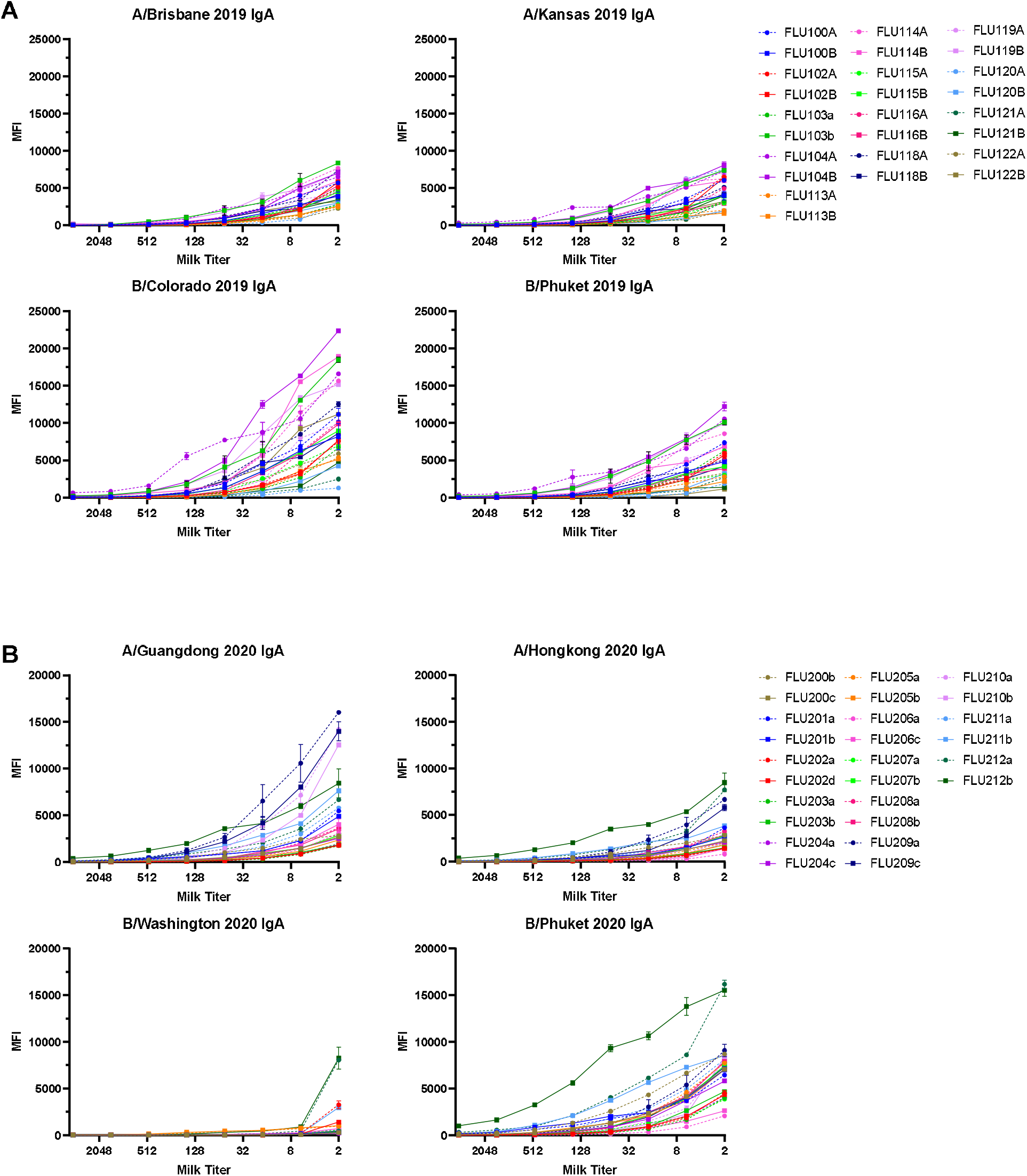
IgA titration curves. (a) 2019-2020 season. (b) 2020-2021 season. Dotted lines indicate pre-vaccine samples, solid lines indicate post-vaccine samples.

**Supplemental Figure 2:**
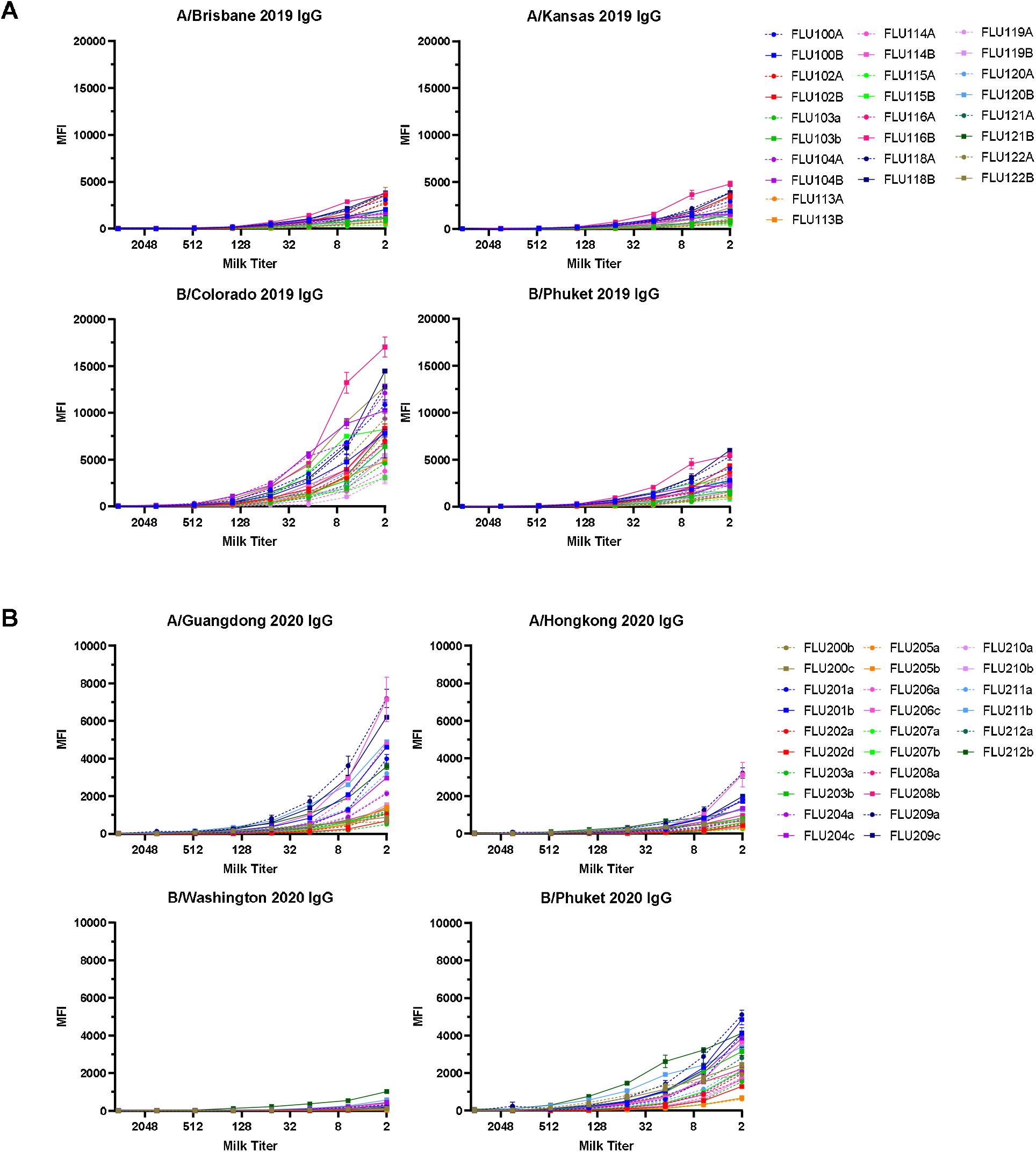
IgG titration curves. (a) 2019-2020 season. (b) 2020-2021 season. Dotted lines indicate pre-vaccine samples, solid lines indicate post-vaccine samples.

**Supplemental Figure 3:**
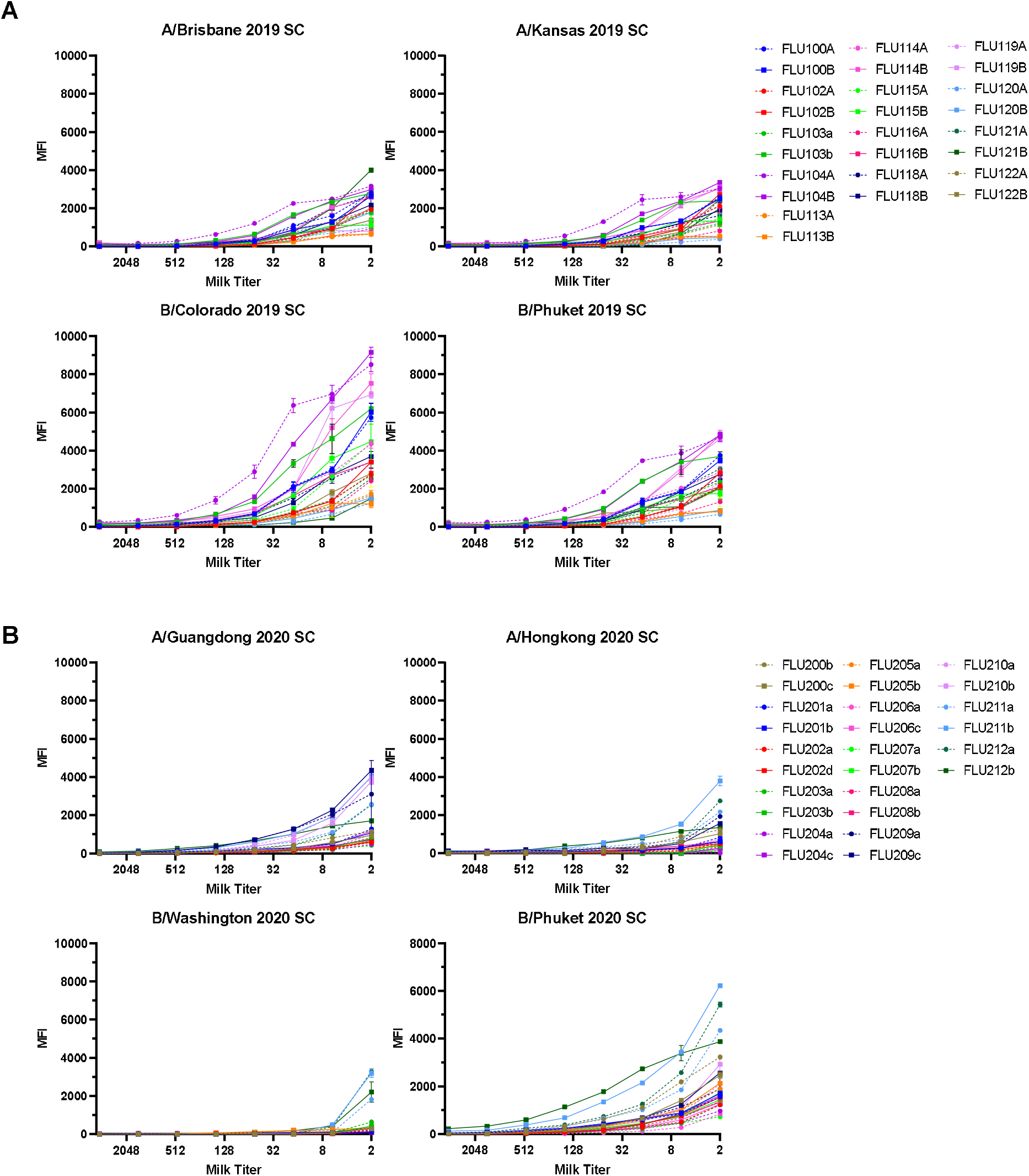
sAb titration curves (anti-SC ELISA). (a) 2019-2020 season. (b) 2020-2021 season. Dotted lines indicate pre-vaccine samples, solid lines indicate post-vaccine samples.

**Supplemental Figure 4:**
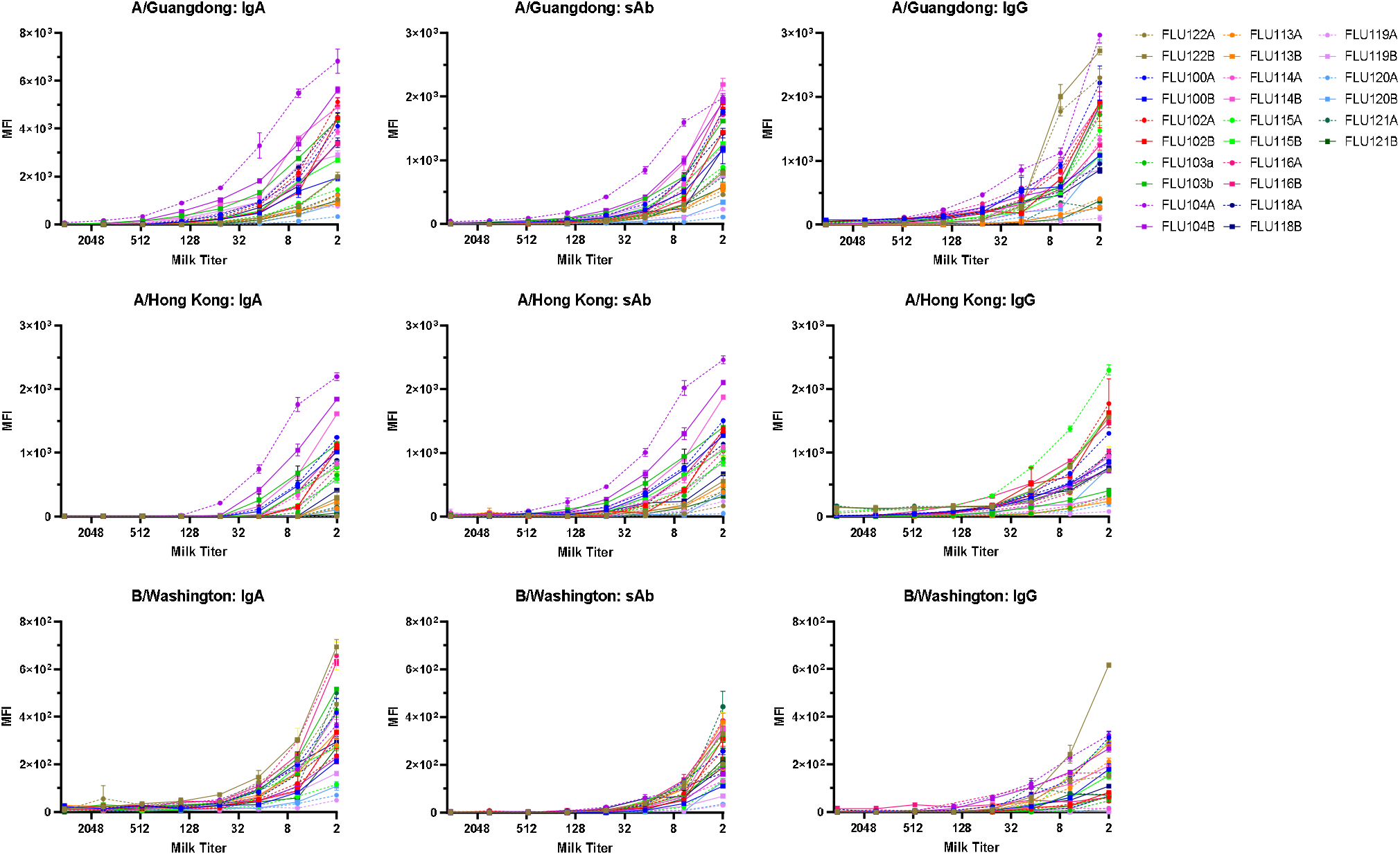
Seasonally-mismatched titration curves.

